# How Large Language Models Can Affect Clinical Reasoning: A Randomized Clinical Trial

**DOI:** 10.64898/2025.12.17.25342338

**Authors:** Mark Levels, Levels, Rounding, Dijksman, Fouarge, Fregin, Steens, Velasco, De Boer, De Bont, Leijenaar, Arif, Findyartini, Greviana, Soemantri, Wildan, Yusuf, Manji, Mbithi, Obungu, Sokwalla, Gmyrek, Berg

## Abstract

**Importance:** LLMs have encoded a vast array of medical knowledge and are being integrated into clinical settings as decision-support tools to improve physician performance across various aspects of care. However, evidence of the impact of LLMs on the clinical reasoning of physicians remains limited.

**Objective:** To evaluate the impact of LLM on core aspects of physicians’ clinical reasoning: diagnostic reasoning, information gathering, and management reasoning in primary care scenarios.

**Design, Setting, and Participants:** We conducted three identical randomized controlled trials (RCTs) in 2024–2025 with 249 physicians in Indonesia, Kenya, and the Netherlands. Participants completed four or five clinical vignettes designed to simulate real-world primary care consultations, with half randomized to have access to ChatGPT-4o.

**Main Outcomes and Measures:** Physician quality of care was evaluated using a rubric based on evidence-based clinical guidelines, scored across nine steps of the clinical reasoning process. Primary outcomes were quality scores for diagnostic reasoning, information gathering, and management. Secondary outcomes were quality per answer, number of answers, and less obvious answers.

**Results:** Access to LLMs enhanced information gathering and management reasoning across all countries. Physicians who were assigned the LLM achieved significantly better quality-of-care scores in diagnostic steps in Indonesia (*b*=7.9%, CI: 4.0% to 11.8%, *p*<.001) and Kenya (*b*= 15.1%, CI: 10.2% to 19.9%, *p*<.001) but not the Netherlands (*b*=1.4%, CI: −1.6% to 4.4%, *p*=1.00). Physicians with LLM access also performed better in investigative steps, in Indonesia (*b*=10.7%, CI: 4.2% to 17.1%, *p*=.004), Kenya (*b*=17.1%, CI: 10.3% to 23.9%, *p*<.001), and the Netherlands (*b*=11.9%, CI: 7.7% to 16.1%, *p*<.001). We also found LLM access affected physicians’ scores in management steps (Indonesia: *b*=15.7%, CI: 8.6% to 22.9%, *p*<.001; Kenya: *b*=27.3%, CI: 19.9% to 34.7% *p*<.001; the Netherlands: *b*=12.3%, CI: 7.1% to 17.5%, *p*<.001). We found that LLM access was less useful in management reasoning for more cognitively demanding cases compared to standard patient cases in Indonesia (*b*=-14.1%, CI: −21.4% to −6.8%, *p*<.001) and Kenya (*b*=-12.1%, CI: −19.6% to −4.6%, *p*=.006).

**Conclusions and Relevance:** In this cross-country randomized control trial, we assessed that access to an LLM had significant positive effects on physicians’ clinical reasoning. The effects we found are promising for the further roll-out of LLMs to supplement physicians in their care tasks. They also suggest that the extent to which LLMs can supplement physicians is context dependent.

**Key Points:** *Question:* To what extent do large language models (LLMs) increase physicians’ quality of diagnostic reasoning, information gathering and management reasoning?

*Findings:* In a randomized clinical trial including 249 physicians in Indonesia, Kenya, and the Netherlands, access to an LLM significantly enhanced clinical reasoning performance in information gathering and management reasoning across all countries, and diagnostic reasoning in Kenya and Indonesia.

*Meaning:* This study shows that the use of an LLM can enhance clinical reasoning of physicians. Further research is needed to effectively understand the augmentation of physician clinical practice.

## Introduction

Large Language Models (LLMs) harbour great potential in augmenting physicians’ clinical reasoning tasks; i.e., constructing differential diagnosis, gathering and processing information, and supporting management reasoning related to tests and treatments is well-documented^1–3^. Although LLMs can assist physicians by augmenting human cognition^4,5^, and specialized LLMs aim to augment differential diagnoses^6^, one randomized clinical trial suggests the impact of LLMs on diagnostic reasoning appears limited^7^.

LLMs have encoded a vast array of medical knowledge to support clinical reasoning^8,9^. Physicians are increasingly exploring the use of LLMs to support diagnostic reasoning, information gathering, management reasoning, and adherence to clinical guidelines ^10,11^. To understand under which conditions and how LLMs can optimally be used to augment physicians’ clinical reasoning, we need to understand how LLMs align with the cognitive and contextual processes that underlie the various stages of clinical reasoning^12^.

We performed identical prospective randomized controlled trials to assess the extent to which having access to an LLM increases physicians’ performance on clinical reasoning in three distinct national contexts: Indonesia, Kenya, and the Netherlands. Participating doctors were presented with sequential information flows on patients’ conditions that mimic real-life clinical settings. Chosen cases simulated primary care scenarios on prevalent medical conditions. We compared the LLMs’ impact on the quality of diagnostic reasoning, information gathering, and management reasoning, as codified in evidence-based, best practice guidelines. Further, we also assessed the role of the cognitive demand of clinical tasks and analysed how LLMs affected quality of reasoning. The objective is not simply to identify the causal effect of LLMs, which has been done. ^1,7^ Rather it is to assess how physicians combined insights from LLMs with their own tacit expert knowledge to arrive at higher quality scores. By conducting comparable RCTs in three countries, we can explore generalizability of LLM impacts across diverse contexts^13,14^.

## Methods

The study was reviewed and approved by review boards of the participating universities (University of Indonesia, Aga Khan University Nairobi, and Maastricht University). Written informed consent was obtained from participants preceding enrolment and randomization. Participants were not compensated for participating in this study. We follow the CONSORT reporting guideline for randomized trials. The study protocol is available in Supplement 1. The study design was preregistered April 17, 2024 at AEA RCT Registry (RCT ID: AEARCTR-0013399).

Participants were recruited via university networks of the medical teaching faculties of the participating universities. We primarily recruited resident physicians in internal or family medicine. This was supplemented by physicians not specialising in internal or family medicine in Kenya (33) and attending physicians specialising in family medicine in the Netherlands (16). Data were collected in August-September 2024 in the Netherlands; November 2024 in Indonesia; and January 2025 in Kenya. 249 physicians (60 in Kenya, 81 in Indonesia and 108 in the Netherlands) completed the study. Sessions were organised in controlled environments: computer labs in Indonesia and the Netherlands, and medical training rooms resembling consultation offices in Kenya. Participants were briefed about the study design and the aim of the research. The study flow is available in eFigure 1 in Supplement 3.

### Clinical Patient Vignettes

Participants were presented with four to five fictional patient vignettes, each measuring quality of care using Peabody’s^15–17^ framework. This framework combines the high internal validity of experiments with the high external validity of surveys to unravel predictors of clinicians’ decision-making^18,19^. Expert physicians from the participating countries collaboratively developed the vignettes to reflect globally prevalent yet varied medical scenarios.

The vignettes quantify the overall quality of care in a single measurement^15^. The vignettes were designed to evaluate quality of clinical reasoning at a granular level and to mimic the sequential and conditional nature of clinical reasoning. To do so, our vignette design included nine steps. We asked physicians to provide open-ended responses on the following topics: 1) giving a first differential diagnosis based on limited information; 2) asking about patient history; 3) giving additional differential diagnoses; 4) conducting physical examinations; 5) giving a third differential diagnosis; 6) additional research or investigations; 7) final differential diagnosis; 8) prescribing medication or treatment and 9) further advice. In between steps participants progressively received additional information about patients’ history and symptoms (after step 2), the outcomes of physical examinations (after step 4) and additional investigations (after step 6).

The vignettes were identical in the three countries. They presented patient conditions related to five different disease groups (cardiovascular, respiratory, musculoskeletal disease, fatigue and infectious disease and rare genetic disorders). Four vignettes had a comparable difficulty level: representing conditions common in all countries with strong evidence-based guidelines, referred to as standard vignettes. A fifth vignette, field tested in the Netherlands and administered in Kenya and Indonesia, was designed to be more cognitively demanding in three main ways. First, the condition was less prevalent in the three countries. Second, the condition is not covered by a single guideline, thus more challenging for physicians to manage. Third, the case was designed to be more complex than appears at first glance.

### Measuring quality of care

Three physician co-authors developed an expert rubric containing a list of necessary answers for all steps, based on national guidelines, evidence-based best practice, and common teaching practices in the three countries. This rubric comprehensively covered the necessary steps required for a correct medication and diagnosis plan. This procedure created a rubric of responses required to effectively diagnose and treat the patient case. We assigned up to 1 point for each item on the rubric. Following Peabody et al^15^, rubric items were weighted to signify importance. More important items were given a higher weight, i.e., 1, compared to 0.5 and 0.33 for less important items. Incorrect or unnecessary answers were not awarded points but also not penalized. Weighting could differ between the countries, depending on the assessment of national experts involved in the design of the patient cases and the rubric. The rubric is available in eTable 1 in Supplement 2.

### Study Design

We conducted three identical parallel group randomized controlled trails in a superiority framework. In three field labs, physicians were fully randomized to either use ChatGPT-4o to assist their clinical reasoning (intervention group) or conduct the clinical reasoning process without support (control group).

The study was conducted using a survey tool (Qualtrics) in which we programmed the stages of the vignette evaluations. For the intervention group, the survey provided integrated access to ChatGPT-4o, with the LLM hard-coded to ensure the same model was used throughout the study. Participants in the control group did not have access to the LLM. The four comparable vignettes were presented in random order. The fifth, more demanding, vignette was always presented last, to impose a maximum cognitive burden. Participants were instructed to conduct anamnesis, draw differential diagnoses, collect additional information, and decide on appropriate treatments. Prior to randomisation, all participants received an instruction sheet on writing high-quality prompting provided in eTable 2 in Supplement 3; they were encouraged to use the LLM if they could. Transcripts of interactions with the LLM were saved. All participants were given a two-hour window for the complete experiment but not forced to complete the task within this time frame. Participants were encouraged to provide the best-quality answers.

### Assessment of quality scores

To assess the quality of clinical reasoning, a panel of expert reviewers blinded to treatment assignment compared participants’ open-ended responses to the items listed in the rubric. Reviewers assessed whether the rubric item was present in participants’ responses. All vignettes were doubly evaluated to reduce the influence of reviewer bias on the outcome of interest.

### Outcome measures

Our primary outcome measure is a quality-of-care score *Q*, defined as:

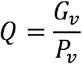

where *G*_*v*_ is the weighted sum of rubric answers given, *P*_*v*_ is the weighted sum of total possible rubric answers, and *v* denotes the vignettes. We generated the quality-of-care score for each step within the vignettes as the average score as assessed by the two reviewers. We combined the scores for the four diagnostic steps (vignette steps 1, 3, 5, and 7) to measure the average quality of diagnostic reasoning. The combined scores for the three information gathering steps (vignette steps 2, 4, and 6) measure the average quality of information gathering. The combined scores for patient management steps (vignette steps 8 and 9) measure the quality of management reasoning.

Secondary outcomes, taken as the average of the reviewers across the outcome: Quality Per Answer, Number of Answers, Less Obvious answers. Each refers to rubric answers that are assessed as present by reviewers per vignette step. For all secondary outcomes: First, we measure the Quality Per Answer as the average rubric weight of present answers. Second, we measure the Number of Answers as the sum of present answers. Third, we measure the Less Obvious Answers as answers that are assessed as present for a small proportion (<25%) of the control group only. The outcome is the sum of Less Obvious Answers.

### Statistical analyses

We predetermined minimal sample sizes of 50 participants with a power analysis on the primary outcome, using Stata software, version 17 (StataCorp). The power analysis demonstrated that there was greater than 80% power to detect a score difference of 4.1%, assuming a two-sided significance level (α) of 0.05 and 5 completed vignettes. The power analysis was clustered at the participant level, incorporating an intraclass correlation coefficient (ICC) of 0.9 and a standard deviation of 5.4%.

Our analyses follow the intention-to-treat principle. Primary analyses were performed at vignette-step levels, standard errors were clustered by participant. All participants completed all vignettes shown to them and were analysed. To estimate intervention effects on the three clinical reasoning outcomes, we used multivariate linear regression models with a conditional likelihood approach. The models conditioned on vignette and vignette-step strata to account for design-related clustering. Covariates included participant gender, clinical experience, and prior exposure to generative AI.

Model assumptions (e.g., normality, homoscedasticity) were verified. Bonferroni corrections were applied to adjust for multiple comparisons and control family-wise type I error (*α*=.05) for all outcome variables. Preplanned sensitivity analyses presented in eTables 3, 4, 5, 6, 7 and 8 in Supplement 3 demonstrate that the estimation of intervention effects was insensitive to alternative model, outcome, and sample specifications.

All statistical analyses were performed using the reghdfe suite of Stata 17, to deal efficiently with fixed effects on multiple levels.

## Results

The study included 249 practicing physicians specialising in internal medicine and family medicine, and physicians with training in internal and family medicine, but pre-residency and specialising in other fields. We had 81 participants in Indonesia, 60 participants in Kenya, and 108 participants in the Netherlands. Table 1 reports on baseline characteristics. Our analyses consisted of three steps. We first compared quality scores between participants who were randomly assigned to use the LLM and those who were not. In the second step, we studied the relevance of the LLM for cases with varying levels of information complexity. Third, we analysed the intervention effects on three secondary outcomes: Quality Per Answer, Number of Answers, and Less Obvious Answers.

**Table 1.**
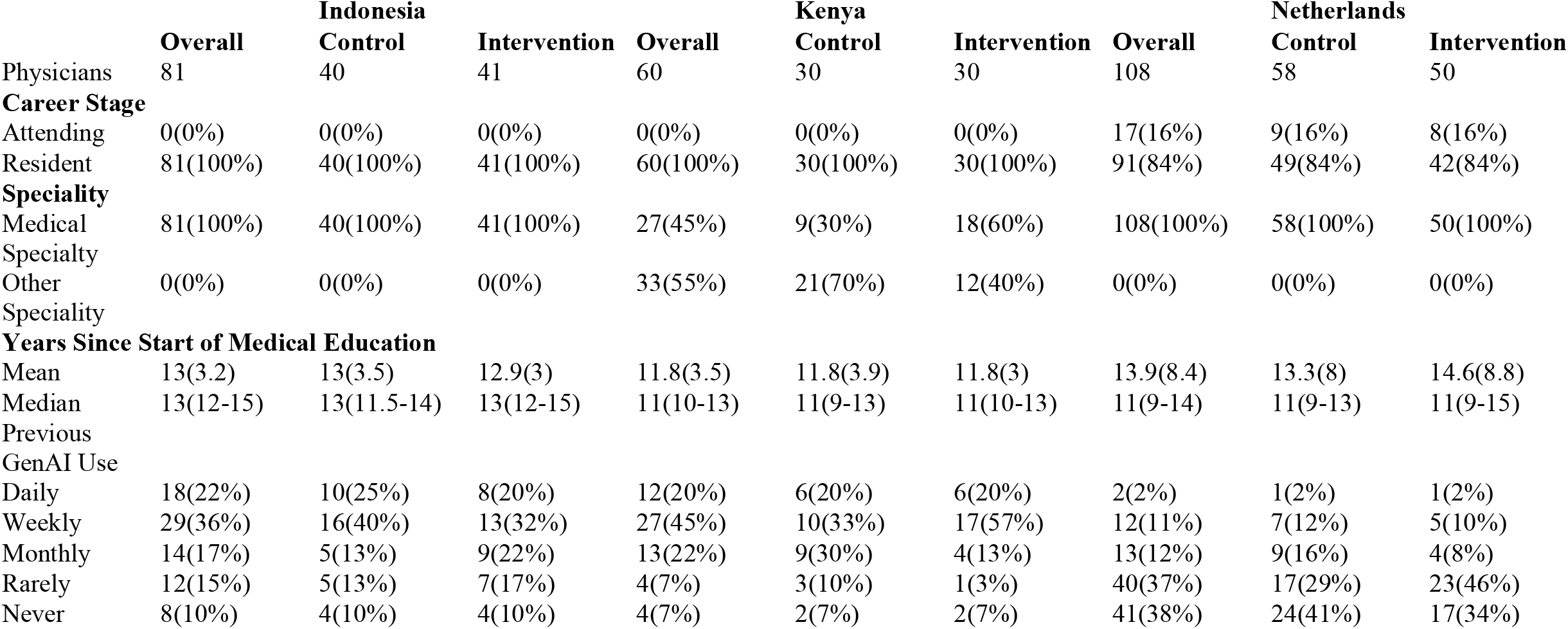
Comparison of Baseline Characteristics between Control and Intervention groups.

### Overall quality of clinical reasoning

In all three countries, the participants randomly assigned to use the LLM achieved higher scores in almost all steps of the clinical reasoning process (Table 2). Physicians who were assigned the LLM achieved significantly better quality-of-care scores in diagnostic steps in Indonesia (*b*=7.9%, CI: 4.0% to 11.8%, *p*<.001) and Kenya (*b*= 15.1%, CI: 10.2% to 19.9%, *p*<.001) but not the Netherlands (*b*=1.4%, CI: −1.6% to 4.4%, *p*=1.00). Physicians with LLM access also perform better in investigative steps, in Indonesia (*b*=10.7%, CI: 4.2% to 17.1%, *p*=.004), Kenya, (*b*=17.1%, CI: 10.3% to 23.9%, *p*<.001), and the Netherlands (*b*=11.9%, CI: 7.7% to 16.1%, *p*<.001). We also find LLM access affected physicians’ scores in management steps (Indonesia: *b*=15.7%, CI: 8.6% to 22.9%, *p*<.001; Kenya: *b*=27.3%, CI: 19.9% to 34.7% *p*<.001; the Netherlands: *b*=12.3%, CI: 7.1%-17.5%, *p*<.001).

**Table 2.**
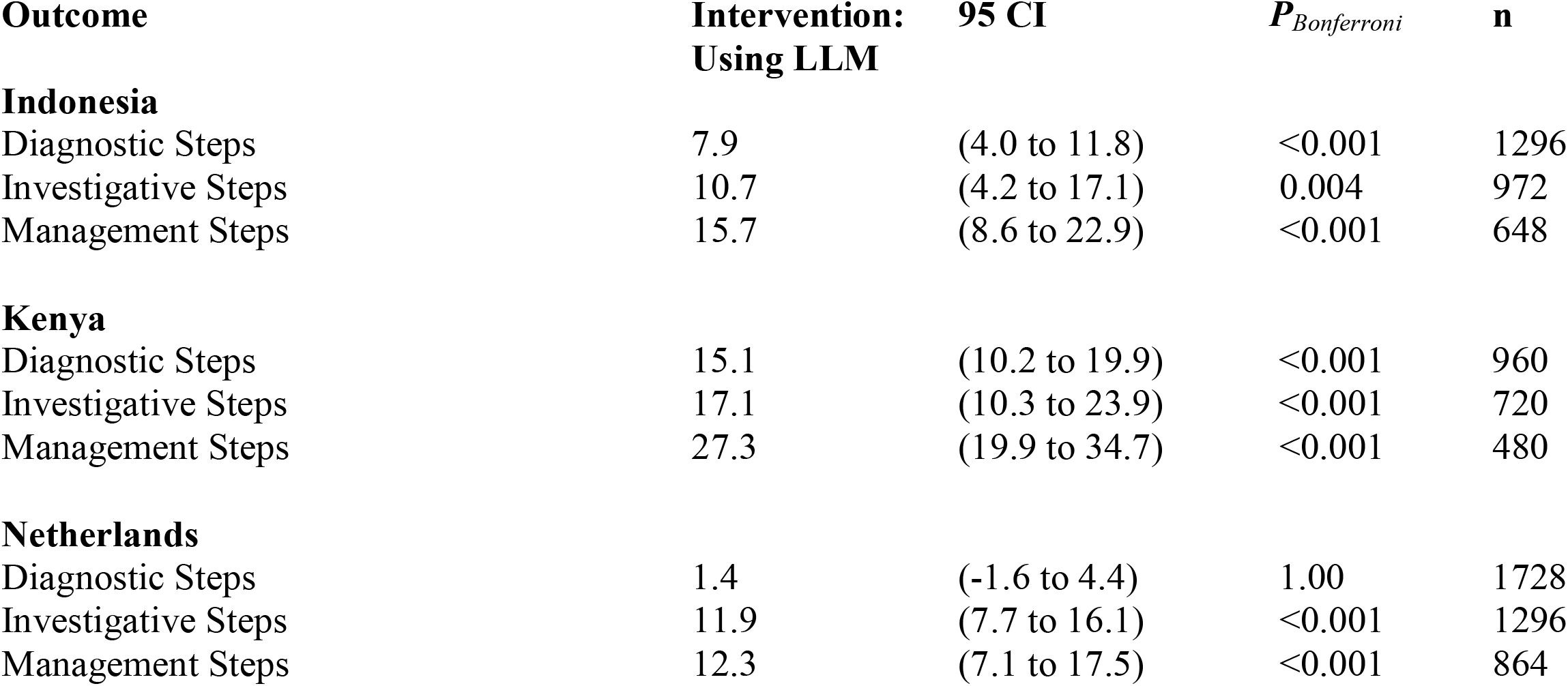
Effect of LLM access on physicians performance on aspects of clinical reasoning.

### Effect of Cognitively Demanding Case

Table 3 demonstrates that physicians on average achieved lower quality-of-care scores for the more cognitively demanding vignette across all steps of the clinical reasoning process in Indonesia and Kenya, with the strongest difference occurring in diagnostic steps in Indonesia (*b*=-25.0%, CI: −29.2% to −20.8%, *p*<.001) and Kenya (*b*=- 32.4%, CI: −37.8% to −27.1%, *p*<.001). In Indonesia, physicians with access to the LLM did not perform significantly better or worse in diagnostic steps and investigative steps and performed worse in management steps (*b*=-14.1%, CI: −21.4% to −6.8%, *p*<.001) when compared to their performance on the standard vignettes. In Kenya we found a positive effect of LLM access for the diagnostic steps when compared to the effect of LLM access on the standard vignettes (*b*=13.2%, CI: 6.6% to 19.8%, *p*<.001), no effect for the investigative steps and, as in Indonesia, a negative effect for the management steps (*b*=-12.1%, CI: −19.6% to −4.6%, *p*=.006).

**Table 3.**
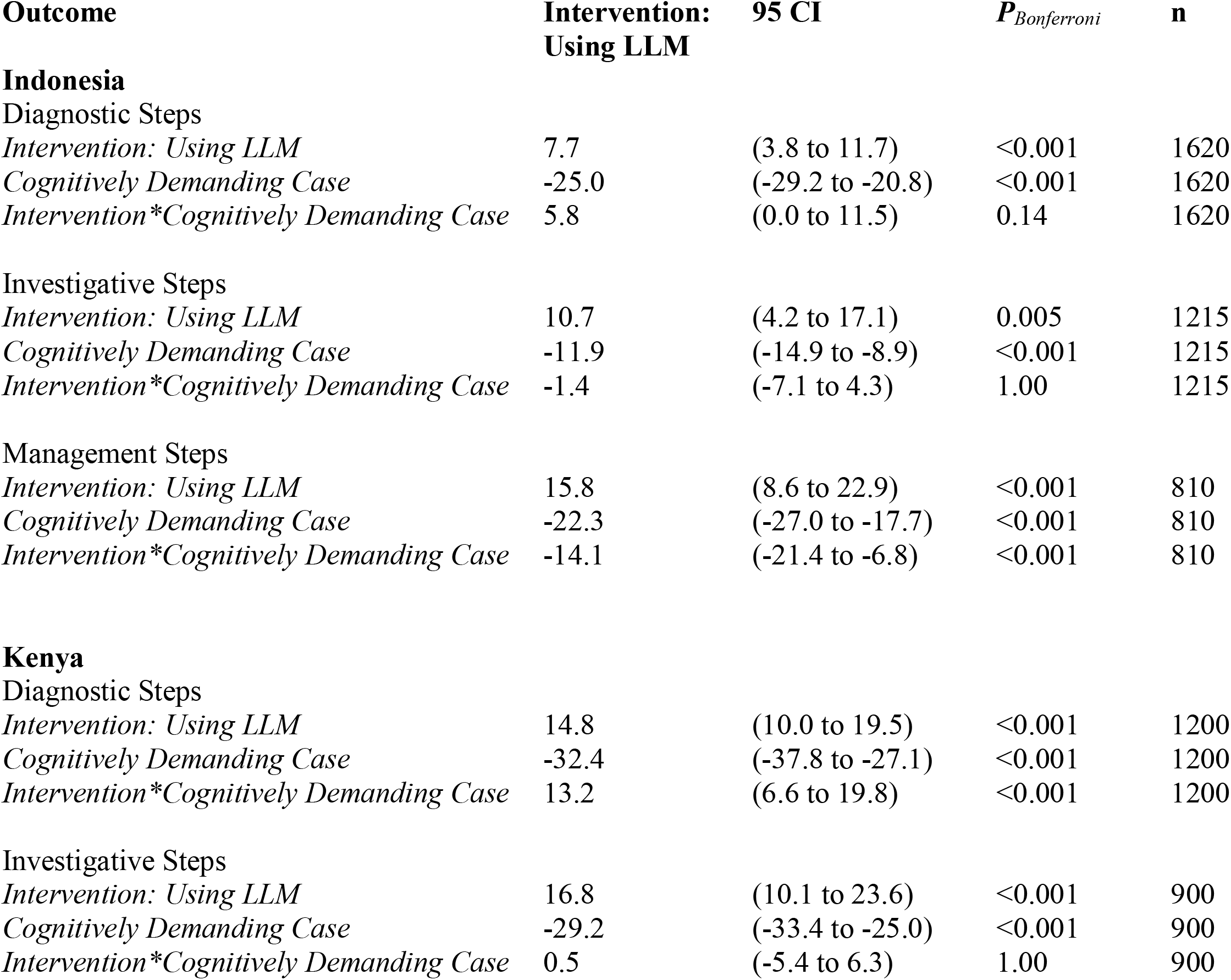
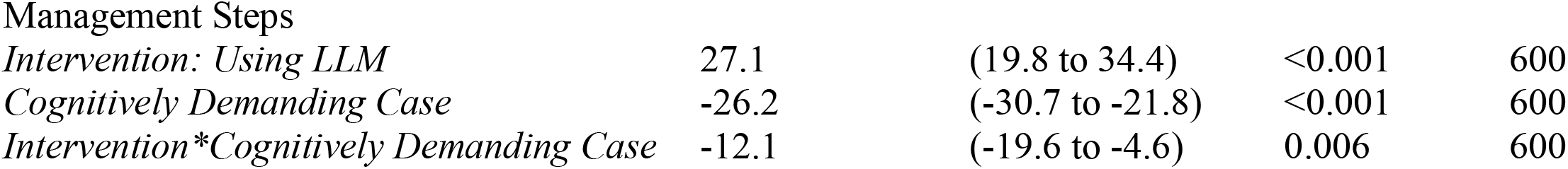

| **Outcome** | **Intervention:**<br>**Using LLM** | **95 CI** | ***P*<sub>Bonferroni</sub>** | **n** |
| --- | --- | --- | --- | --- |
| **Indonesia** |  |  |  |  |
| Diagnostic Steps |  |  |  |  |
| *Intervention: Using LLM* | 7.7 | (3.8 to 11.7) | <0.001 | 1620 |
| *Cognitively Demanding Case* | -25.0 | (-29.2 to -20.8) | <0.001 | 1620 |
| *Intervention\*Cognitively Demanding Case* | 5.8 | (0.0 to 11.5) | 0.14 | 1620 |
| Investigative Steps |  |  |  |  |
| *Intervention: Using LLM* | 10.7 | (4.2 to 17.1) | 0.005 | 1215 |
| *Cognitively Demanding Case* | -11.9 | (-14.9 to -8.9) | <0.001 | 1215 |
| *Intervention\*Cognitively Demanding Case* | -1.4 | (-7.1 to 4.3) | 1.00 | 1215 |
| Management Steps |  |  |  |  |
| *Intervention: Using LLM* | 15.8 | (8.6 to 22.9) | <0.001 | 810 |
| *Cognitively Demanding Case* | -22.3 | (-27.0 to -17.7) | <0.001 | 810 |
| *Intervention\*Cognitively Demanding Case* | -14.1 | (-21.4 to -6.8) | <0.001 | 810 |
| **Kenya** |  |  |  |  |
| Diagnostic Steps |  |  |  |  |
| *Intervention: Using LLM* | 14.8 | (10.0 to 19.5) | <0.001 | 1200 |
| *Cognitively Demanding Case* | -32.4 | (-37.8 to -27.1) | <0.001 | 1200 |
| *Intervention\*Cognitively Demanding Case* | 13.2 | (6.6 to 19.8) | <0.001 | 1200 |
| Investigative Steps |  |  |  |  |
| *Intervention: Using LLM* | 16.8 | (10.1 to 23.6) | <0.001 | 900 |
| *Cognitively Demanding Case* | -29.2 | (-33.4 to -25.0) | <0.001 | 900 |
| *Intervention\*Cognitively Demanding Case* | 0.5 | (-5.4 to 6.3) | 1.00 | 900 |
| Management Steps |  |  |  |  |
| *Intervention: Using LLM* | 27.1 | (19.8 to 34.4) | <0.001 | 600 |
| *Cognitively Demanding Case* | -26.2 | (-30.7 to -21.8) | <0.001 | 600 |
| *Intervention\*Cognitively Demanding Case* | -12.1 | (-19.6 to -4.6) | 0.006 | 600 |

### Secondary outcomes

Table 4 demonstrates intervention effects on three alternative outcomes. Having access to an LLM did not increase the probability of giving more high-quality answers in Indonesia, Kenya, or the Netherlands. In all countries, participants with LLM access had more rubric items assessed as present by reviewers than those that did not. LLM access also increased the probability of giving a less obvious answer.

**Table 4.**

| **Outcome** | **Intervention:**<br>**Using LLM** | **95 CI** | ***P*<sub>Bonferroni</sub>** | **n** |
| --- | --- | --- | --- | --- |
| **Indonesia** |  |  |  |  |
| Quality Per Answer | -0.008 | (-0.018 to 0.003) | 0.16 | 2588 |
| Number of Answers | 0.491 | (0.256 to 0.725) | <0.001 | 2916 |
| Less Obvious Answers | 0.294 | (0.194 to 0.394) | <0.001 | 2916 |
| **Kenya** |  |  |  |  |
| Quality Per Answer | -0.002 | (-0.012 to 0.009) | 0.77 | 2028 |
| Number of Answers | 0.800 | (0.544 to 1.055) | <0.001 | 2160 |
| Less Obvious Answers | 0.259 | (0.184 to 0.334) | <0.001 | 2160 |
| **Netherlands** |  |  |  |  |
| Quality Per Answer | -0.006 | (-0.015 to 0.002) | 0.11 | 3720 |
| Number of Answers | 0.359 | (0.213 to 0.506) | <0.001 | 3888 |
| Less Obvious Answers | 0.146 | (0.106 to 0.185) | <0.001 | 3888 |

### Assessment validation

The Cohen κ value for rubric items assessed as present between the two initial reviewers was 0.72 (Indonesia: 0.68; Kenya: 0.68; Netherlands: 0.79) indicating substantial agreement between reviewers^20^. One-way random-effects intraclass correlation coefficient (ICC) for our primary outcome was 0.905 (95% CI: .902 to .909; *p*<.001), indicating excellent agreement in composite scores between graders^21^. To account for any disagreements between reviewers, we recruited a third reviewer to assess the disagreements and decide the outcome. The third reviewers were expert physicians from the three countries. Sensitivity analyses presented in eTable 4 and eTable 7 demonstrate that the analyses are insensitive to reviewer evaluation methods and disagreements.

## Discussion

Various studies suggest LLMs can achieve above human performance on clinical reasoning tasks, leading some to suggest that physicians could be replaced^1,22,23^. However, the low error tolerance and ethical need for transparent reasoning make full replacement unlikely^24^. The core question is to understand under which circumstances physicians can best work with LLMs.

Our findings show that LLM access can enhance clinical reasoning in diagnostic reasoning, information gathering and management reasoning. This largely confirms prior findings from U.S.-based studies^1,7^ for the African, Asian and European context, though with some important contextually dependent differences. We found LLM usage improved diagnostic reasoning in Indonesia and Kenya, but not in the Netherlands. As the average score on diagnostic steps was already very high for Dutch physicians without LLM access, this points towards potential ceiling effects that diminish returns to LLM use for those that already score well. Physicians with LLM access performed better at investigative tasks and patient management tasks in all three countries. Our data suggest that LLM access is less useful in cognitively demanding settings for management reasoning when compared to the standard cases. On the contrary, we found that LLM access was more beneficial for diagnostic reasoning in Kenya, compared to the standard vignettes. We also found that providing access to an LLM did not lead to better answers in the standard vignettes, but to a higher number of good answers, and to a higher likelihood of finding rare but necessary alternatives. In one country (Kenya), we observed that physicians with LLM access were more likely to come up with irrelevant alternatives.

This sheds light on the way in which LLM use can affect clinical reasoning. Physicians rely on both intuitive and analytical approaches to clinical reasoning and use them adaptively, depending on the case complexity and other relevant contexts^25,26^. Intuitive clinical reasoning can help to generate early diagnostic hypotheses based on limited information and compare them to patterns stored in long-term memory^27^. However, it is prone to cognitive biases^28^ that reduce the quality of clinical reasoning^29^. Analytical approaches to clinical reasoning – while cognitively demanding and data hungry – can effectively counteract such biases^30^. As LLMs essentially process information analytically, they may improve clinical reasoning by producing information that challenges potentially erroneous priors^26^. This is demonstrated by the higher likelihood of physician’s with LLM access providing less obvious differential diagnosis, investigate strategies, or management actions. A potential downside to this may be an increased risk of overdiagnoses.

However, LLMs can also cause clinical reasoning errors, and provide inappropriate and poor quality answers in diagnostic tasks^31,32^. One issue is that LLM outputs are biased by the type of medical data they are trained on^33^. Particularly when information on patients’ disease history is biased, LLMs are equally likely as human doctors to reach false conclusions^34^. Also, hallucination is an intrinsic feature of LLM models^34^ that seriously hampers LLM’s ability to reliably process and communicate medical information^35^.LLMs can provide misleading or biased information, but even accurate outputs warrant caution. They may induce cognitive biases, such as conjunction errors or base rate neglect. Future research must assess their impact on diagnostic error.

## Limitations

This study has limitations. First, the results we present are lab results. Our conclusions have a high internal validity, but the external validity of laboratory experiments is notably debatable. There is ample reason to be cautious about LLMs’ merit outside of experimental lab conditions^36^. Therefore, randomised controlled trails of LLM use by physicians in clinical settings need to be conducted to test whether our results hold. Second, we used a commercially available LLM that was trained on general data. However, this choice reflects the current reality, where access to more specialized, medically trained LLMs remains limited. In future studies, the use of medically trained LLMs is recommended^6^.

## Conclusions

Participants who had access to an LLM performed better at most diagnostic reasoning, information gathering and management reasoning tasks in a randomized clinical trial in Indonesia, Kenya, and the Netherlands. Further, participants with LLM access provided more answers and less obvious alternatives, but not necessarily better answers on average. The effects on more cognitively demanding cases are ambiguous. The extent to which LLMs can improve clinical reasoning thus depends on physicians’ meta-cognitive ability to critically assess their own reasoning and the information and clinical reasoning provided by the LLM^37^.

## Supporting information

Supplement 1 - study protocol

Supplement 2 - eTable 1 - Rubric Score and Vignette information and questions v20250716

Supplement 3 - Online Content (eFigure1, eTables,2,3,4,5,6,7,8)

Supplement 4 - Data Sharing Statement 20250724

## Data Availability

All data produced in the present study are available upon reasonable request to the authors

## ARTICLE INFORMATION

### Author Contributions

Levels and Rounding had full access to all of the data in the study and take responsibility for the integrity of the data and the accuracy of the data analysis. Levels and Rounding contributed equally to this work.

### Concept and design

All authors.

### Acquisition of data

Levels, Rounding, Steens, Dijksman, Arif, Findyartini, Leijenaar, De Bont, Mbithi, Manji, Soemantri, Sokwalla, Cals.

### Analysis or interpretation of data

Levels, Rounding, Steens, Dijksman, Fregin, De Bont, Leijenaar, Findyartini, Manji, Wildan.

### Drafting of the manuscript

Levels, Rounding, Fregin.

### Critical review of the manuscript for important intellectual content

All authors. Statistical analysis: Levels, Rounding, Dijksman, Steens

### Obtained funding

Levels, Rounding, Berg, Gmyrek, Velasco. Administrative, technical, or material support: Dijksman, Steens. Conflict of Interest Disclosures: No other disclosures were reported.

### Data Sharing Statement

See Supplement 4.

## Supplement 1

Trial Protocol

## Supplement 2

eTable 1: Vignettes and expert rubric containing a list of necessary answers for all steps, based on the national guidelines and common teaching practices in the three countries.

## Supplement 3

eFigure 1: Study Flow Diagram by Country

eTable 2: Prompting Sheet

eTable 3: Sensitivity Analysis 1 – Alternate Model Specification

eTable 4: Sensitivity Analysis 2 – Alternative Primary Outcome after third round of assessment

eTable 5: Sensitivity Analysis 3 – Primary Outcomes removing third diagnostic step

eTable 6: Sensitivity Analysis 4 – Primary model and outcomes at vignette case level

eTable 7: Sensitivity Analysis 5 – Primary model and outcomes at vignette case level removing those with more than 10% disagreement

eTable 8: Sensitivity Analysis 6 – Primary model and outcomes for Kenya split between Internal Medicine Specialists and Physicians specialising in other fields

### Supplement 4

Data Sharing Statement

