## Supplement 1 - study protocol for "How Large Language Models Can Affect Clinical Reasoning: A Randomized Clinical Trial"

**Title stating the trial design, population, and interventions**

**Structured summary of trial design and methods, including items from the World Health Organization Trial Registration Data Set**

A parallel group randomized controlled trial using a superiority framework. Clinical vignettes were used to assess the impact of a large language model on the clinical reasoning of physicians. Physicians were asked to complete a set of ‘clinical vignettes’ hypothetical but viable patient scenarios in a computer lab. Each vignette consisted of 9 steps: 1) giving a first differential diagnosis based on limited information; 2) asking about patient history; 3) giving additional differential diagnoses; 4) conducting physical examinations; 5) giving a third differential diagnosis; 6) additional research or investigations; 7) final differential diagnosis; 8) prescribing medication or treatment and 9) further advice. In each step, physicians were asked to provide answers in an online Qualtrics survey. Half of the physicians were randomly assigned an AI assistant. The AI assistant was be an interface developed by UM researchers that will use the OpenAI API key and GPT-4. The other half did not receive the AI assistant.

**Trial Registration:**

AEA Registry: https://www.socialscienceregistry.org/trials/13399

The Big Unknown: A Journey into Generative AI's Transformative Effect on Professions, starting with Medical Practitioners

RCT ID: AEARCTR-0013399

Initial registration date: April 17, 2024

Initial registration date is when the trial was registered.

It corresponds to when the registration was submitted to the Registry to be reviewed for publication.

First published: April 25, 2024, 11:44 AM EDT

First published corresponds to when the trial was first made public on the Registry after being reviewed.

**Study Design**

Study Type: Interventional (Clinical Trial)

Intervention Model: Parallel group trial design with an allocation of 1:1 in a superiority framework

Allocation: Simple randomization without stratification generated by the Qualtrics randomizer function and the Evenly Present Elements option

Settings: Computer access points in universities in Kenya, Netherlands and Indonesia

Eligibility criteria for participants: We recruited resident physicians in internal and family medicine in Indonesia, Kenya, and the Netherlands. We also recruited residents in their first year of study in other specialties and post-internship, pre-residency physicians (referred to as Senior House Officers).

Actual enrolment: Indonesia:81; Kenya:60; Netherlands: 108

Intervention and comparator: The intervention is access to ChatGPT 4o via the OpenAI API key provided in a Qualtrics environment.

Blinding: all participants were aware of the intentions of the study prior to randomisation.

Blinding Graders: The grading of responses was performed by graders who were blinded to participant identity and the control and intervention condition.

Actual Study Start Date:

Actual Primary Completion Date:

Actual Study Completion Date:

**Study Arms**

| **Arm** | **Intervention / Treatment** |
| --- | --- |
| Active Comparator: GPT-4  Group will be given access to GPT-4 via an accessed via a custom built interface integrated into the Qualtrics survey tool via an iFrame | Other: GPT-4 OpenAI's GPT-4 large language model with chat interface. |
| No Intervention: no additional resources |  |

**Outcome Measures**

In each country, two independent reviewers randomly selected from a pool will grade physicians’ answers in each step against a rubric. The pool sizes were 4 in Kenya, 8 in Indonesia and 11 in the Netherlands. Reviewers were asked to assess whether an expert answer could be found in the response entered into the Qualtrics environment.

To account for disagreements in the assessment of rubric answers between the primary two graders, we recruited a third reviewer in each country. The reviewers were more experienced physician who had been engaged in the experimental set-up and design. This reviewer was presented with the physician response and the rubric answer in which the reviewers were in disagreement. Therefore, after the third review process all rubric answers had two physicians in agreement on the response.

Primary outcomes are a percent correct score generated through the assessment of physician responses against a rubric. The primary outcome variable is generated as a percent correct score, dividing the weighted total sum of rubric items assessed as present by the total number of rubric items possibly at the level at which the score is generated, i.e. vignette or vignette step level. Outcomes will be generated for different parts of the reasoning process: Vignette Steps 1,3,5,7 are diagnostic;2,4,6 are investigative, 8&9 are patient management. Primary outcomes are a mean of the scores generated by the two reviewers.

Secondary outcomes are generated as the average weighted score of responses assessed as present, the total number of items assessed as present, and rubric items that are less frequently assessed as present for the control group. We use a cut off of 75%, preplanned sensitivity analyses are conducted to assess the impact of the cut off on results.

One concern for the outcome measures is the third vignette step, which is asked as: Please provide any additional diagnoses. Due to the wording of this step, many participants chose to leave this blank. To adjust this, we append the scores of rubric items assessed as present in Step 1 to the Step 3 scores if that rubric item appears in the rubric for both steps in order to generate a more accurate percentage correct score. This procedure doubly weights responses provided in Step 3. To assess the effect of this change on the vignette score and the difference between intervention and control, pre-planned sensitivity analyses will be run with this question removed from the analysis.

**Power analysis**

A two mean power analysis was conducted using Stata/SE 17.0^[1]^ to determine the appropriate sample size for detecting meaningful effects in the randomized controlled trial. Using vignette parameters derived from the literature (Peabody et al, 2000), including a mean vignette score of 71 and standard deviation of 5.4, a two-means clustered power analysis was implemented using Stata 17, treating each participant as a cluster and each vignette as an observation. With an assumed intra-cluster correlation of 0.9 and targeting a power of 0.8, the analysis suggested that a minimum of 50 participants would be sufficient to detect a lower-bound effect size of 4.8%, based on literature estimates of AI's impact on physician performance papers (Bien et al., 2018; Han et al., 2020; Jain et al., 2021; Jussupow et al., 2021)

Statistical Analysis:

Descriptive analysis will be performed using Stata/SE 17.0. Initial descriptives results comparing the AI assisted vs non-assisted group, percentages and numbers will be shown for categorical variables and means, standard deviations, medians and interquartile ranges will be shown for continuous variables.

The analysis will be conducted at the question level for each country separately using ordinary least squares regressions, clustering for standard errors at the participant level and including vignette and question covariates to control for variance related to each level. Primary outcomes will be run on the different stages of the clinical reasoning process: management, investigative, and diagnostic reasoning. Heterogeneity analyses will be run across experience levels, for gender, and pre-experiment generative AI use. A more cognitively challenging case will be analysed separately. Primary and secondary outcomes will all be continuous and analysed separately.

Preplanned sensitivity analyses will be conducted using a mixed effects model with intercepts for participants, vignettes and vignette questions. Further pre-planned sensitivity analyses will be conducted restricting the data sample to:

- Rubric items found in all three rubrics, to account for country specific rubric effects
- The data where a third reviewer has adjudicated any disagreements on the assessment of rubric items in the first review round
- Removing vignettes where there is more than a 10% disagreement between the first and second reviewer.
- Removing the third question

1. power twomeans 71, sd(5.4) k1(25) k2(25) m1(5) m2(5) power(0.8) rho(0.9)
