## Supplement 3 - Online Content (eFigure1, eTables,2,3,4,5,6,7,8) for "How Large Language Models Can Affect Clinical Reasoning: A Randomized Clinical Trial"

**eFigure 1: Study Flow Diagram by Country**

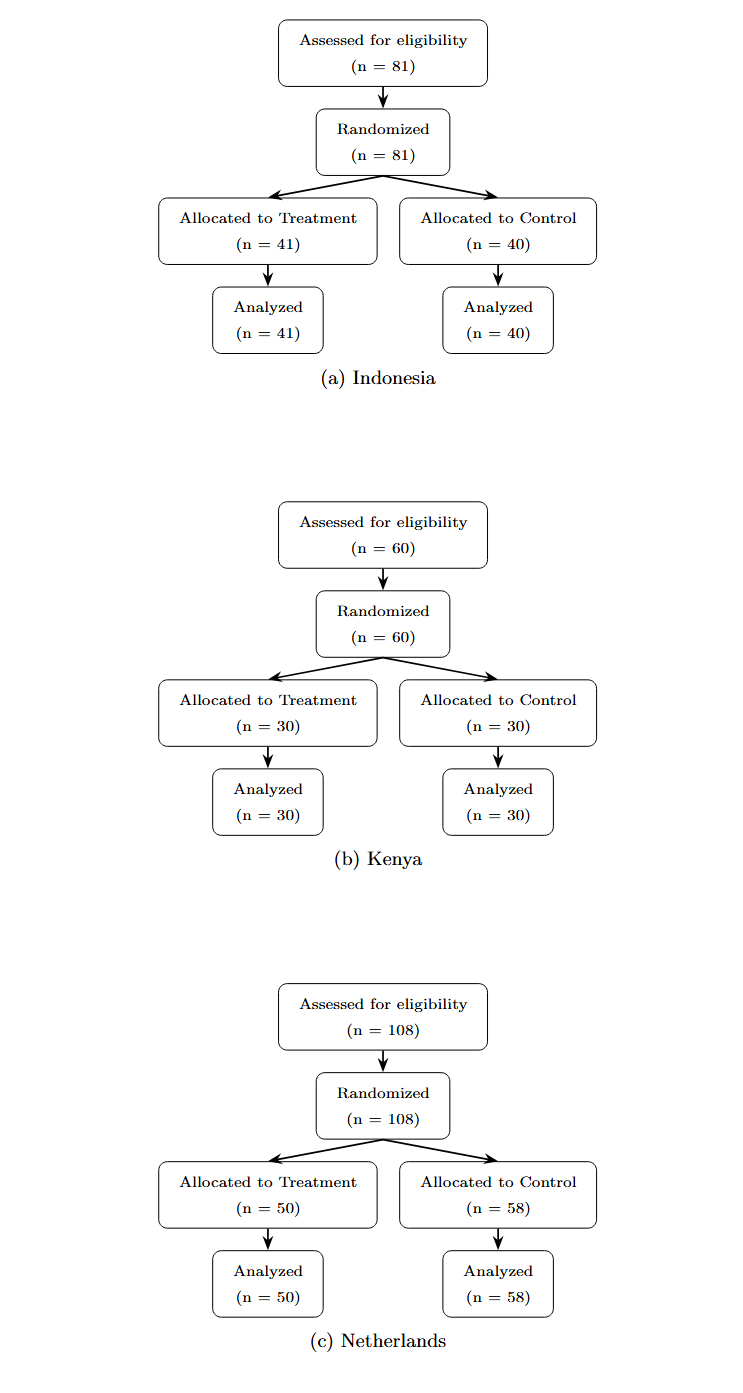

eTable 2: Prompting Sheet

eTable 3: Sensitivity Analysis 1 – mixed effects

eTable 4: Sensitivity Analysis 2 – 3^rd^ reviewer

eTable 5: Sensitivity Analysis 3 – No Step 3

eTable 6: Sensitivity Analysis 4 – Vignette Level

eTable 7: Sensitivity Analysis 5 – Vignette Level with cases with higher than 10% disagreement removed

eTable 8: Descriptive information about the construction of the vignettes (including prevalence rates and information complexity)

eTable 2 – Prompting instructions

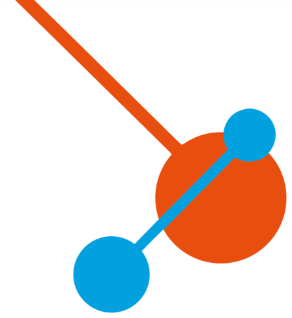
**Briefing Prompting**

**AI in Primary Care**

**Huisartsenopleiding x ROA/ SBE**

**An Event for General Practitioners**

Dear Participants,

Thank you for joining this experiment. In this study, we aim to evaluate the impact of AI on your diagnostic reasoning. During the session, you are allowed to use a chatbot to help you.

**Tips on how to interact with the chatbot:**

- Ask clear and specific questions.
- Include how you want the chatbot to present its answer (e.g., create a list that contains…).
- Specify how long the answer should be; this will influence the level of detail you will receive in the chatbot’s answer.
- Give the chatbot one task or question at a time.
- You can copy and paste from the chatbot.
- The safeguards of the chatbot don’t always allow you to get answers to specific questions, such as asking for medication advice, unless you prompt it creatively by e.g. describing your role and medical experience.

**Tips on how NOT to interact with the chatbot:**

- Avoid ambiguity and leading questions.
- Don't overload with information but specify the steps  
  you require the chatbot to take.
- Don’t repeat your questions if the answer was not right but build on your previous prompts.
- Don't expect perfection, always be critical of its answers and correct the chatbot when it makes a mistake.
- And very important: never feed the chatbot any personal data.

There are no wrong answers; please respond to the best of your knowledge. For the quality of the research, we also request that you refrain from using your phone or any other form of communication during the experiment. Additionally, please remain seated at your computer and avoid using any functionalities other than the browser tab provided for the experiment. 

Thank you for your cooperation!

eTable 3: Sensitivity Analysis 1 – Alternate Model Specification (Mixed Effects Model)

| **Outcome** | **Intervention: Using LLM** | **95 CI** | ***P****_Bonferroni_* | **n** |
| --- | --- | --- | --- | --- |
| **Indonesia** |  |  |  |  |
| Diagnostic Steps | 7.9 | (4.0 to 11.8) | <0.001 | 1296 |
| Investigative Steps | 10.7 | (4.3 to 17.1) | 0.003 | 972 |
| Management Steps | 15.7 | (8.7 to 22.7) | <0.001 | 648 |
| **Kenya** |  |  |  |  |
| Diagnostic Steps | 15.1 | (10.6 to 19.5) | <0.001 | 960 |
| Investigative Steps | 17.1 | (10.8 to 23.4) | <0.001 | 720 |
| Management Steps | 27.3 | (20.6 to 34.0) | <0.001 | 480 |
| **Netherlands** |  |  |  |  |
| Diagnostic Steps | 1.4 | (-1.6 to 4.3) | 1.00 | 1728 |
| Investigative Steps | 11.9 | (7.8 to 16.0) | <0.001 | 1296 |
| Management Steps | 12.3 | (7.4 to 17.2) | <0.001 | 864 |

eTable 4: Sensitivity Analysis 2 – Alternative Primary Outcome after third round of assessment

| **Outcome** | **Intervention: Using LLM** | **95 CI** | ***P****_Bonferroni_* | **n** |
| --- | --- | --- | --- | --- |
| **Indonesia** |  |  |  |  |
| Diagnostic Steps | 9.0 | (5.0 to 13.0) | <0.001 | 1296 |
| Investigative Steps | 11.7 | (4.7 to 18.6) | 0.004 | 972 |
| Management Steps | 16.9 | (9.4 to 24.5) | <0.001 | 648 |
| **Kenya** |  |  |  |  |
| Diagnostic Steps | 15.8 | (10.8 to 20.8) | <0.001 | 960 |
| Investigative Steps | 18.1 | (11.3 to 24.9) | <0.001 | 720 |
| Management Steps | 27.7 | (20.0 to 35.4) | <0.001 | 480 |
| **Netherlands** |  |  |  |  |
| Diagnostic Steps | 2.1 | (-1.0 to 5.2) | 0.54 | 1728 |
| Investigative Steps | 13.6 | (9.2 to 18.1) | <0.001 | 1296 |
| Management Steps | 12.7 | (7.1 to 18.4) | <0.001 | 864 |

eTable 5: Sensitivity Analysis 3 – Primary Outcomes removing third diagnostic step

| **Outcome** | **Intervention: Using LLM** | **95 CI** | ***P****_Bonferroni_* | **n** |
| --- | --- | --- | --- | --- |
| **Indonesia** |  |  |  |  |
| Diagnostic Steps | 7.3 | (3.3 to 11.3) | 0.002 | 972 |
| Investigative Steps | 10.7 | (4.2 to 17.1) | 0.004 | 972 |
| Management Steps | 15.7 | (8.6 to 22.9) | <0.001 | 648 |
| **Kenya** |  |  |  |  |
| Diagnostic Steps | 15.2 | (10.2 to 20.2) | <0.001 | 720 |
| Investigative Steps | 17.1 | (10.3 to 23.9) | <0.001 | 720 |
| Management Steps | 27.3 | (19.9 to 34.7) | <0.001 | 480 |
| **Netherlands** |  |  |  |  |
| Diagnostic Steps | 0.5 | (-2.5 to 3.5) | 1.00 | 1296 |
| Investigative Steps | 11.9 | (7.7 to 16.1) | <0.001 | 1296 |
| Management Steps | 12.3 | (7.1 to 17.5) | <0.001 | 864 |

eTable 6: Sensitivity Analysis 4 – Primary model and outcomes at vignette case level

| **Outcome** | **Intervention: Using LLM** | **95 CI** | ***P****_Bonferroni_* | **n** |
| --- | --- | --- | --- | --- |
| **Indonesia** |  |  |  |  |
| Diagnostic Steps | 7.8 | (4.0 to 11.6) | <0.001 | 324 |
| Investigative Steps | 10.7 | (4.3 to 17.2) | 0.004 | 324 |
| Management Steps | 15.6 | (8.9 to 22.3) | <0.001 | 324 |
| **Kenya** |  |  |  |  |
| Diagnostic Steps | 14.3 | (9.5 to 19.1) | <0.001 | 240 |
| Investigative Steps | 17.1 | (10.2 to 23.9) | <0.001 | 240 |
| Management Steps | 24.7 | (17.8 to 31.6) | <0.001 | 240 |
| **Netherlands** |  |  |  |  |
| Diagnostic Steps | 1.4 | (-1.6 to 4.4) | 1.00 | 432 |
| Investigative Steps | 11.6 | (7.4 to 15.8) | <0.001 | 432 |
| Management Steps | 11.1 | (6.1 to 16.2) | <0.001 | 432 |

eTable 7: Sensitivity Analysis 5 – Primary model and outcomes at vignette case level removing those with more than 10% disagreement

| **Outcome** | **Intervention: Using LLM** | **95 CI** | ***P****_Bonferroni_* | **n** |
| --- | --- | --- | --- | --- |
| **Indonesia** |  |  |  |  |
| Diagnostic Steps | 7.8 | (3.7 to 11.8) | <0.001 | 270 |
| Investigative Steps | 11.1 | (4.4 to 17.7) | 0.004 | 270 |
| Management Steps | 13.9 | (6.9 to 20.9) | <0.001 | 270 |
| **Kenya** |  |  |  |  |
| Diagnostic Steps | 14.2 | (9.3 to 19.2) | <0.001 | 211 |
| Investigative Steps | 16.9 | (9.6 to 24.3) | <0.001 | 211 |
| Management Steps | 23.8 | (16.2 to 31.4) | <0.001 | 211 |
| **Netherlands** |  |  |  |  |
| Diagnostic Steps | 1.7 | (-1.4 to 4.7) | 0.85 | 392 |
| Investigative Steps | 11.6 | (7.3 to 16.0) | <0.001 | 392 |
| Management Steps | 12.1 | (6.6 to 17.5) | <0.001 | 392 |

eTable 8: Sensitivity Analysis 5 – Primary model and outcomes for Kenya split between Internal Medicine Specialists and Physicians specialising in other fields

| **Outcome** | **Intervention: Using LLM** | **95 CI** | ***P****_Bonferroni_* | **n** |
| --- | --- | --- | --- | --- |
| **All** |  |  |  |  |
| Diagnostic Steps | 15.1 | (10.2 to 19.9) | <0.001 | 960 |
| Investigative Steps | 17.1 | (10.3 to 23.9) | <0.001 | 720 |
| Management Steps | 27.3 | (19.9 to 34.7) | <0.001 | 480 |
| **Internal Medicine Specialists** |  |  |  |  |
| Diagnostic Steps | 6.6 | (-0.2 to 13.5) | 0.17 | 432 |
| Investigative Steps | 3.4 | (-5.8 to 12.5) | 1.00 | 324 |
| Management Steps | 18.1 | (7.7 to 28.4) | 0.004 | 216 |
| **Not Specialising in Internal Medicine** |  |  |  |  |
| Diagnostic Steps | 18.2 | (10.8 to 25.5) | <0.001 | 528 |
| Investigative Steps | 22.0 | (13.0 to 31.0) | <0.001 | 396 |
| Management Steps | 29.5 | (19.1 to 39.8) | <0.001 | 264 |
