## Supplement 4 - Data Sharing Statement 20250724 for "How Large Language Models Can Affect Clinical Reasoning: A Randomized Clinical Trial"

**Data types and scope:**
De‑identified participant‑level data
(demographics, survey response, graded vignette response, outcomes), analysis scripts (Stata17).
**Documentation:**
Data dictionary in CSV; annotated Stata scripts.
**Repository and access:**
All materials archived in DataverseNL (https://dataverse.nl), restricted access.
**Timing:**
Available upon article publication.
**Use permissions:**
Available for reproducibility or collaborative academic research, taking into account privacy constraints. Usage only under legal framework.
**Contact:**
Sanne Steens.
